# Effect of vaccination and of prior infection on infectiousness of vaccine breakthrough infections and reinfections

**DOI:** 10.1101/2021.07.28.21261086

**Authors:** Laith J. Abu-Raddad, Hiam Chemaitelly, Houssein H. Ayoub, Patrick Tang, Peter Coyle, Mohammad R. Hasan, Hadi M. Yassine, Fatiha M. Benslimane, Hebah A. Al Khatib, Zaina Al Kanaani, Einas Al Kuwari, Andrew Jeremijenko, Anvar Hassan Kaleeckal, Ali Nizar Latif, Riyazuddin Mohammad Shaik, Hanan F. Abdul Rahim, Gheyath K. Nasrallah, Mohamed Ghaith Al Kuwari, Adeel A. Butt, Hamad Eid Al Romaihi, Abdullatif Al Khal, Mohamed H. Al-Thani, Roberto Bertollini

## Abstract

SARS-CoV-2 breakthrough infections in vaccinated individuals and in those who had a prior infection have been observed globally, but the transmission potential of these infections is unknown. The RT-qPCR cycle threshold (Ct) value is inversely correlated with viral load and culturable virus. Here, we investigated differences in RT-qPCR Ct values across Qatar’s national cohorts of primary infections, reinfections, BNT162b2 (Pfizer-BioNTech) breakthrough infections, and mRNA-1273 (Moderna) breakthrough infections. Through matched-cohort analyses of the randomly diagnosed infections, the mean Ct value was higher in all cohorts of breakthrough infections compared to the cohort of primary infections in unvaccinated individuals. The Ct value was 1.3 (95% CI: 0.9-1.8) cycles higher for BNT162b2 breakthrough infections, 3.2 (95% CI: 1.8-4.5) cycles higher for mRNA-1273 breakthrough infections, and 4.0 (95% CI: 3.4-4.6) cycles higher for reinfections in unvaccinated individuals. Assuming a linear relationship between viral load and infectiousness, these differences imply that breakthrough infections are at least 50% less infectious than primary infections in unvaccinated individuals. Public health benefits of vaccination may have been underestimated, as COVID-19 vaccines not only protect against acquisition of infection, but also appear to protect against transmission of infection.

## Introduction

Coronavirus Disease 2019 (COVID-19) vaccines have demonstrated protection against the severe acute respiratory syndrome coronavirus 2 (SARS-CoV-2)^1–3^. However, the efficacy against acquisition of infection is imperfect in that vaccines have an efficacy *VE*_*S*_ < 100%, particularly against variants of concern^4,5^. Breakthrough infections in vaccinated individuals have been documented in various countries^4–6^, but the transmission potential of these infections is poorly understood. It is conceivable that vaccinated persons who acquire the infection may be less infectious than unvaccinated persons who acquire the infection, as the vaccine-primed immune response may attenuate the natural history of infection and reduce viral replication, leading to lower viral load and faster infection clearance^7^. There is evidence that seem to support this hypothesis for SARS-CoV-2 infection^8,9^. Therefore, COVID-19 vaccines may not only be efficacious against acquisition of infection (*VE*_*S*_), but also against transmission of infection^7^, thereby adding additional efficacy for each vaccine, denoted as *VE*_*I*_ and defined as the proportional reduction in infectiousness among those infected, but vaccinated, compared to those infected but unvaccinated^7^.

Leveraging the national, federated databases that have captured all SARS-CoV-2 vaccinations and polymerase chain reaction (PCR) testing in Qatar since the start of the epidemic (Methods), we investigated the effect of vaccination on infectiousness by comparing SARS-CoV-2 real time (quantitative) reverse transcription-PCR (RT-qPCR) cycle threshold (Ct) values of individuals infected and fully vaccinated with the values of those infected and unvaccinated. The RT-qPCR Ct value is a measure of the inverse of viral load and correlates strongly with culturable virus^10^; thus, it can be used as a proxy of SARS-CoV-2 infectiousness^10^. We also investigated the effect of prior infection on infectiousness at reinfection by comparing the RT-qPCR Ct values among those reinfected with SARS-CoV-2 with the values among those in primary infection.

These comparisons were implemented utilizing: i) the national cohort of all 384,452 RT-qPCR-confirmed primary infections since epidemic onset (February 28, 2020) until the end of the study (July 11, 2021; Figure 1A); ii) the national cohort of all 1,695 RT-qPCR-confirmed reinfections during the same period (Figure 1B); iii) the national cohort of all 898,648 individuals vaccinated with the BNT162b2^1^ (Pfizer-BioNTech) vaccine since the first recorded vaccination in Qatar on December 16, 2020 until the end of the study (July 11, 2021; Figure 1C); and iv) the national cohort of all 468,872 vaccinated individuals with the mRNA-1273^2^ (Moderna) vaccine during the same period (Figure 1D).

**Figure 1.**
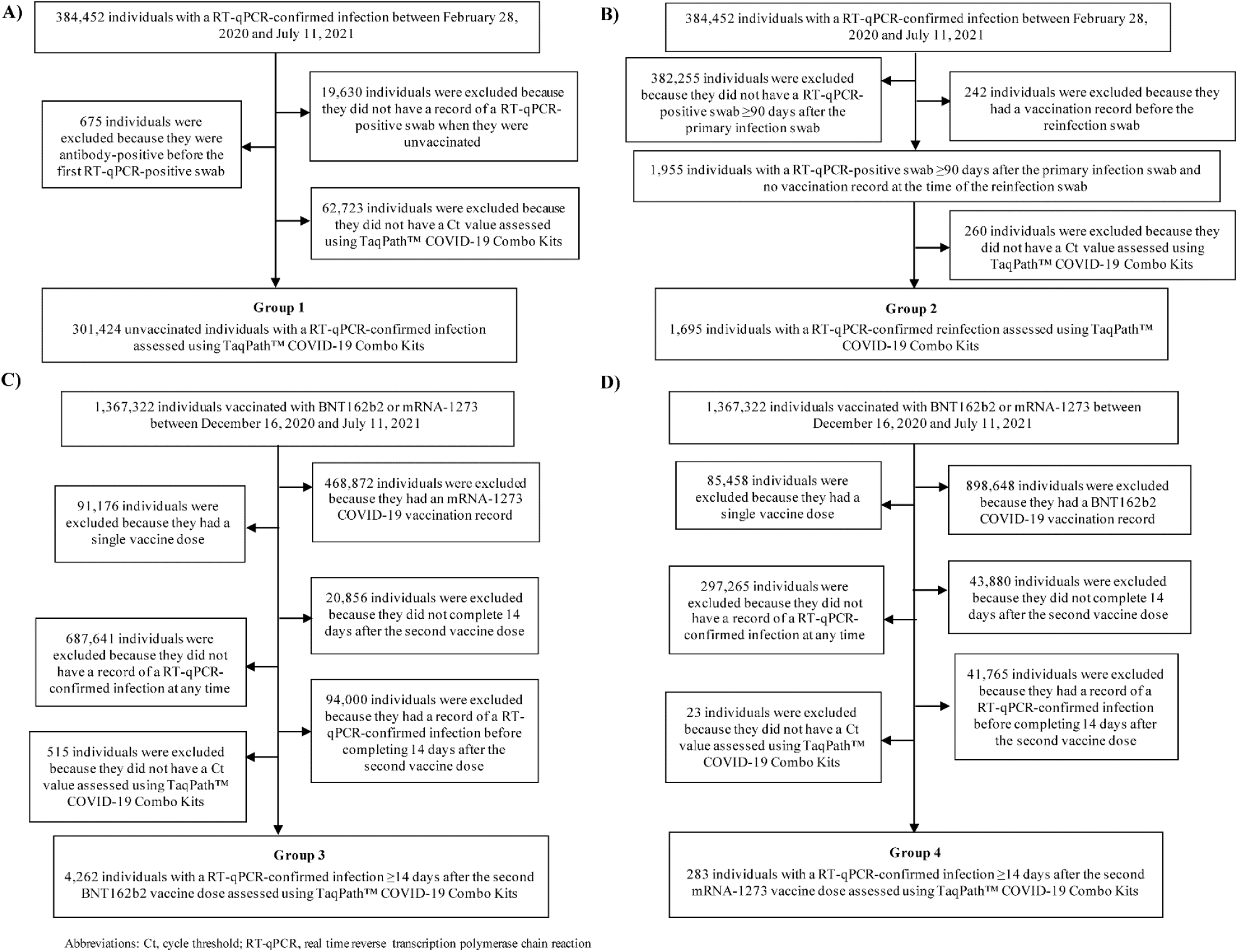
Flowchart illustrating the selection of the cohorts of primary infections in unvaccinated individuals, reinfections in unvaccinated individuals, BNT162b2-vaccine breakthrough infections, and mRNA-1273-vaccine breakthrough infections.

The BNT162b2 and mRNA-1273 vaccines have been the vaccines of choice in the national immunization campaign in Qatar^4,5,11,12^. In total, there has been 4,777 breakthrough infections in those fully vaccinated with BNT162b2 (0.61% of those fully vaccinated; Figure 1C) and 306 mRNA-1273 breakthrough infections in those fully vaccinated with mRNA-1273 (0.09% of those fully vaccinated; Figure 1D). Of note that mass immunization started with the BNT162b2 vaccine, and the mRNA-1273 vaccine was introduced only several weeks later.

Primary infection was defined as the first RT-qPCR-positive test for a given individual. Reinfection was defined as the first RT-qPCR-positive test that occurred ≥90 days after the primary infection^13–16^. Breakthrough infection in a vaccinated individual was defined as an RT-qPCR-positive test 14 or more days after the individual received the second vaccine dose, conditional on this RT-qPCR-positive test being the first ever positive for this individual.

## Results

### Study populations

Figure 1 shows the process for identifying eligible primary infections, reinfections, BNT162b2 breakthrough infections, and mRNA-1273 breakthrough infections. Figure 2 schematizes the six pairwise comparisons conducted between these cohorts of infections, after matching in a 1:1 ratio by sex, 10-year age group, reason for RT-qPCR testing, and calendar week of the RT-qPCR test, to control for differences in biology by sex and age, as well as exposure risk^17,18^ and variant exposure^4,19,20^ (Methods).

**Figure 2.**
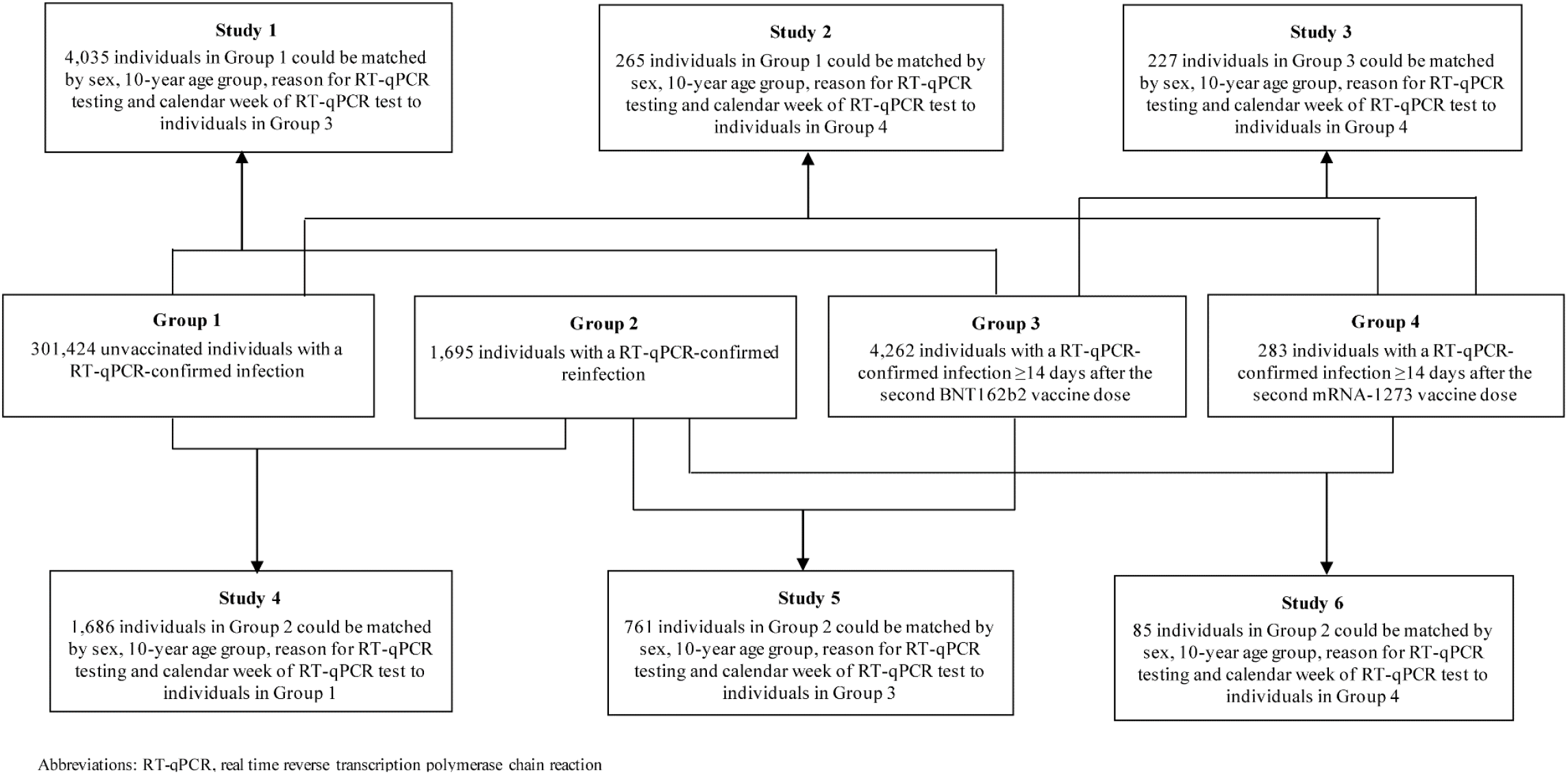
Schematic diagram showing the process of formulating the six pairwise comparisons between the cohorts of primary infections in unvaccinated individuals, reinfections in unvaccinated individuals, BNT162b2-vaccine breakthrough infections, and mRNA-1273-vaccine breakthrough infections, after 1:1 matching by sex, 10-year age group, reason for RT-qPCR testing, and calendar week of the RT-qPCR test.

Supplementary Tables 1 and 2 show demographic characteristics of the study populations in the six pairwise comparisons. Scatter plots of the distribution of RT-qPCR Ct values in each comparison were generated for all RT-qPCR-confirmed infections, regardless of the reason for the RT-qPCR testing (Supplementary Figure 1), only for the randomly diagnosed (asymptomatic) infections (Figure 3; Methods), and only for the symptomatic infections (Supplementary Figure 2; Methods).

**Figure 3.**
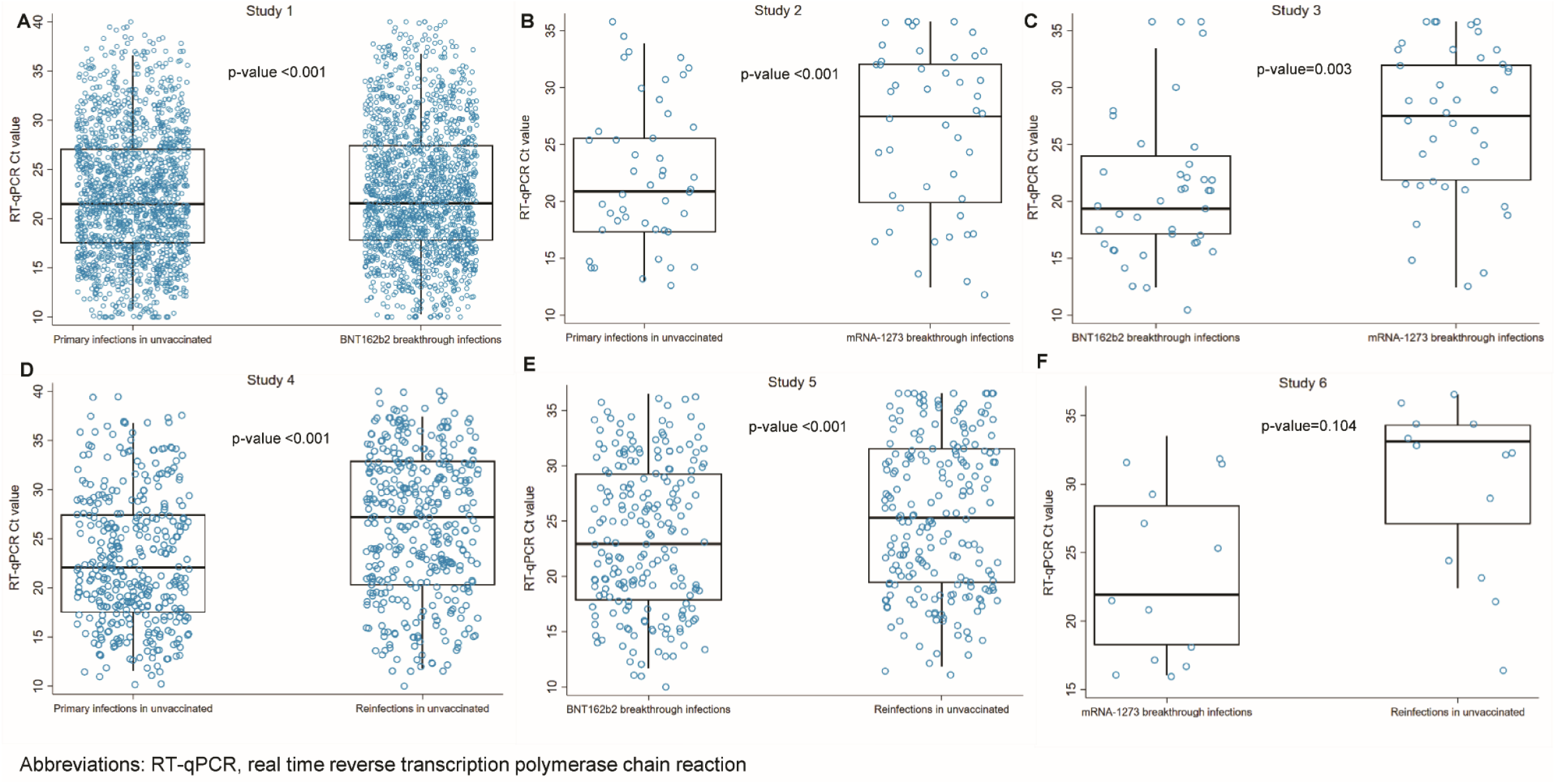
RT-qPCR Ct values in the randomly diagnosed (asymptomatic) SARS-CoV-2 infections. Distribution of these Ct values in the six pairwise comparisons between primary infections in unvaccinated individuals, reinfections in unvaccinated individuals, BNT162b2-vaccine breakthrough infections, and mRNA-1273-vaccine breakthrough infections. A randomly diagnosed infection was defined as an RT-qPCR-positive test conducted with no prior reason to suspect infection and no reported presence of symptoms compatible with a respiratory tract infection. That is, the RT-qPCR test was conducted as part of a survey (random testing campaigns), for routine healthcare testing, for pre-travel requirement, or at port of entry upon arrival in Qatar. Boxplots center lines indicate the median Ct values, box limits indicate the 25% and 75% quartiles, and whiskers indicate maximum and minimum observations within 1.5 of interquartile range.

### Differences in RT-qPCR Ct values in all confirmed infections

In the comparisons including all RT-qPCR-confirmed infections, regardless of the reason for the RT-qPCR testing, the mean RT-qPCR Ct value was higher in all cohorts of breakthrough infections compared to the cohort of primary infections in unvaccinated individuals (Table 1 and Supplementary Figure 1). The Ct value was 1.0 (95% confidence interval (CI): 0.7-1.2) cycles higher for BNT162b2 breakthrough infections, 3.5 (95% CI: 2.4-4.6) cycles higher for mRNA-1273 breakthrough infections, and 3.7 (95% CI: 3.4-4.2) cycles higher for reinfections in unvaccinated individuals. The Ct value was 3.1 (95% CI: 1.9-4.3) cycles higher in mRNA-1273 breakthrough infections than in BNT162b2 breakthrough infections. Compared to reinfections in unvaccinated individuals, the Ct value was 2.0 (95% CI: 0.2-3.9) cycles lower in mRNA-1273 breakthrough infections and 1.7 (95% CI: 1.1-2.4) cycles lower in BNT162b2 breakthrough infections. All differences in Ct values were statistically significant with p-values ≤0.03.

**Table 1.**
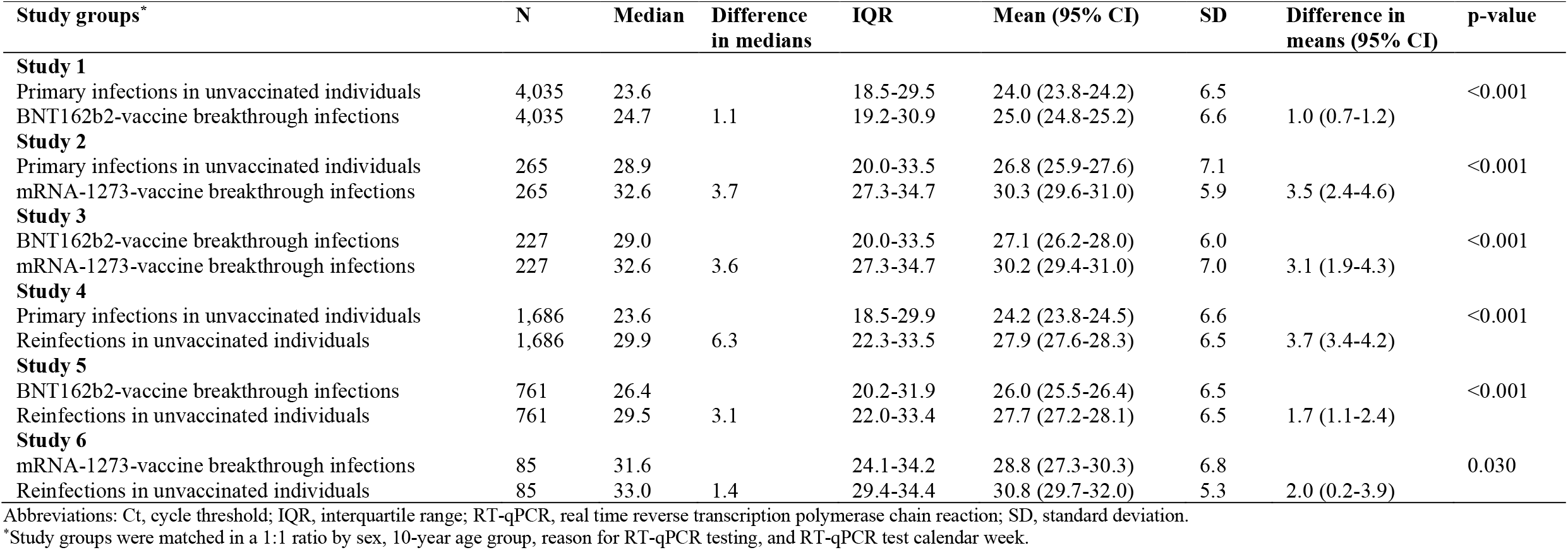
RT-qPCR Ct values, including all confirmed infections, regardless of the reason for the RT-qPCR testing, in the six pairwise comparisons between primary infections in unvaccinated individuals, reinfections in unvaccinated individuals, BNT162b2-vaccine breakthrough infections, and mRNA-1273-vaccine breakthrough infections.

| Study groups* | N | Median | Difference in medians | IQR | Mean (95% CI) | SD | Difference in means (95% CI) | p-value |
| --- | --- | --- | --- | --- | --- | --- | --- | --- |
| **Study 1** |  |  |  |  |  |  |  |  |
| Primary infections in unvaccinated individuals | 4,035 | 23.6 |  | 18.5-29.5 | 24.0 (23.8-24.2) | 6.5 |  | <0.001 |
| BNT162b2-vaccine breakthrough infections | 4,035 | 24.7 | 1.1 | 19.2-30.9 | 25.0 (24.8-25.2) | 6.6 | 1.0 (0.7-1.2) |  |
| **Study 2** |  |  |  |  |  |  |  |  |
| Primary infections in unvaccinated individuals | 265 | 28.9 |  | 20.0-33.5 | 26.8 (25.9-27.6) | 7.1 |  | <0.001 |
| mRNA-1273-vaccine breakthrough infections | 265 | 32.6 | 3.7 | 27.3-34.7 | 30.3 (29.6-31.0) | 5.9 | 3.5 (2.4-4.6) |  |
| **Study 3** |  |  |  |  |  |  |  |  |
| BNT162b2-vaccine breakthrough infections | 227 | 29.0 |  | 20.0-33.5 | 27.1 (26.2-28.0) | 6.0 |  | <0.001 |
| mRNA-1273-vaccine breakthrough infections | 227 | 32.6 | 3.6 | 27.3-34.7 | 30.2 (29.4-31.0) | 7.0 | 3.1 (1.9-4.3) |  |
| **Study 4** |  |  |  |  |  |  |  |  |
| Primary infections in unvaccinated individuals | 1,686 | 23.6 |  | 18.5-29.9 | 24.2 (23.8-24.5) | 6.6 |  | <0.001 |
| Reinfections in unvaccinated individuals | 1,686 | 29.9 | 6.3 | 22.3-33.5 | 27.9 (27.6-28.3) | 6.5 | 3.7 (3.4-4.2) |  |
| **Study 5** |  |  |  |  |  |  |  |  |
| BNT162b2-vaccine breakthrough infections | 761 | 26.4 |  | 20.2-31.9 | 26.0 (25.5-26.4) | 6.5 |  | <0.001 |
| Reinfections in unvaccinated individuals | 761 | 29.5 | 3.1 | 22.0-33.4 | 27.7 (27.2-28.1) | 6.5 | 1.7 (1.1-2.4) |  |
| **Study 6** |  |  |  |  |  |  |  |  |
| mRNA-1273-vaccine breakthrough infections | 85 | 31.6 |  | 24.1-34.2 | 28.8 (27.3-30.3) | 6.8 |  | 0.030 |
| Reinfections in unvaccinated individuals | 85 | 33.0 | 1.4 | 29.4-34.4 | 30.8 (29.7-32.0) | 5.3 | 2.0 (0.2-3.9) |  |

Abbreviations: Ct, cycle threshold; IQR, interquartile range; RT-qPCR, real time reverse transcription polymerase chain reaction; SD, standard deviation.
\*Study groups were matched in a 1:1 ratio by sex, 10-year age group, reason for RT-qPCR testing, and RT-qPCR test calendar week.

### Differences in RT-qPCR Ct values in randomly diagnosed infections

In the comparisons including only the randomly diagnosed (asymptomatic) infections (Methods), which are perhaps most representative for differences between these cohorts of infections, given the random diagnosis, the mean RT-qPCR Ct value was also higher in all cohorts of breakthrough infections compared to the cohort of primary infections in unvaccinated individuals (Table 2 and Figure 3). The Ct value was 1.3 (95% CI: 0.9-1.8) cycles higher for BNT162b2 breakthrough infections, 3.2 (95% CI: 1.8-4.5) cycles higher for mRNA-1273 breakthrough infections, and 4.0 (95% CI: 3.4-4.6) cycles higher for reinfections in unvaccinated individuals. The Ct value was 2.3 (95% CI: 0.8-3.7) cycles higher in mRNA-1273 breakthrough infections than in BNT162b2 breakthrough infections. Compared to reinfections in unvaccinated individuals, the Ct value was 2.0 (95% CI: 1.1-2.8) cycles lower in BNT162b2 breakthrough infections and 1.7 (95% CI: (−0.4-3.8) cycles lower in mRNA-1273 breakthrough infections. All differences in Ct values were statistically significant with p-values ≤0.003 except for the comparison between reinfections and mRNA-1273 breakthrough infections; likely a consequence of the small number of breakthrough infections that have been documented among those vaccinated with the mRNA-1273 vaccine in Qatar.

**Table 2.**
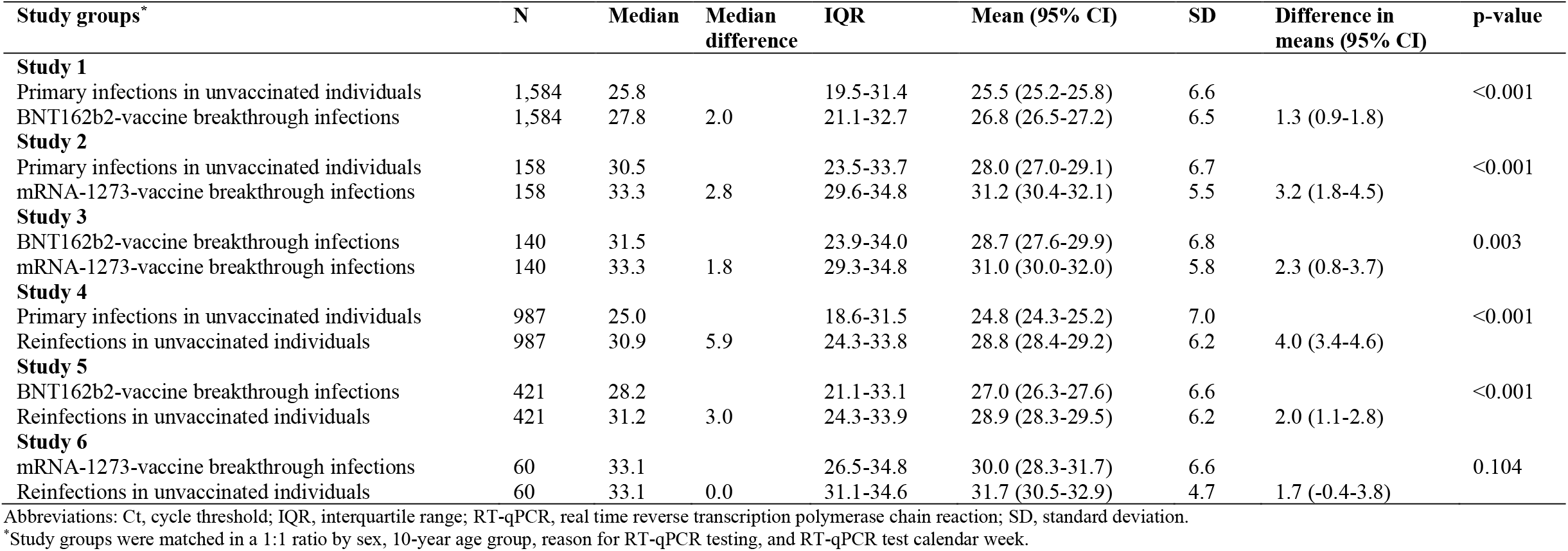
RT-qPCR Ct values, including only the randomly diagnosed (asymptomatic) infections, in the six pairwise comparisons between primary infections in unvaccinated individuals, reinfections in unvaccinated individuals, BNT162b2-vaccine breakthrough infections, and mRNA-1273-vaccine breakthrough infections. A randomly diagnosed infection is defined as an RT-qPCR-positive test conducted with no prior reason to suspect infection and no reported presence of symptoms compatible with a respiratory tract infection.

### Differences in RT-qPCR Ct values in symptomatic infections

In the comparisons including only the symptomatic infections (Methods), the mean RT-qPCR Ct value was also higher in all cohorts of breakthrough infections compared to the cohort of primary infections in unvaccinated individuals, but the difference was smaller for BNT162b2 breakthrough infections (Table 3 and Supplementary Figure 2). The Ct value was 0.2 (95% CI: - 0.2-0.6) cycles higher for BNT162b2 breakthrough infections, 4.9 (95% CI: 2.4-7.4) cycles higher for mRNA-1273 breakthrough infections, and 3.8 (95% CI: 2.8-4.7) cycles higher for reinfections in unvaccinated individuals. The Ct value was 5.3 (95% CI: 2.5-8.2) cycles higher in mRNA-1273 breakthrough infections than in BNT162b2 breakthrough infections. Compared to reinfections in unvaccinated individuals, the Ct value was 1.8 (95% CI: 0.6-3.1) cycles lower in BNT162b2 breakthrough infections and 6.4 (95% CI: 1.3-11.6) cycles lower in mRNA-1273 breakthrough infections. All differences in Ct values were statistically significant with p-values ≤0.016 except for the comparison between primary infections and BNT162b2 breakthrough infections. Notably, very few *symptomatic* mRNA-1273 breakthrough infections have been documented in Qatar (Table 3).

**Table 3.** RT-qPCR Ct values, including only the symptomatic infections, in the six pairwise comparisons between primary infections in unvaccinated individuals, reinfections in unvaccinated individuals, BNT162b2-vaccine breakthrough infections, and mRNA-1273-vaccine breakthrough infections.

| Study groups* | N | Median | Median difference | IQR | Mean (95% CI) | SD | Difference in means (95% CI) | p-value |
| --- | --- | --- | --- | --- | --- | --- | --- | --- |
| **Study 1** |  |  |  |  |  |  |  |  |
| Primary infections in unvaccinated individuals | 1,566 | 21.5 |  | 17.5-31.0 | 22.5 (22.2-22.8) | 6.0 |  | 0.335 |
| BNT162b2-vaccine breakthrough infections | 1,566 | 21.5 | 0.0 | 17.8-27.5 | 22.7 (22.4-23.0) | 6.0 | 0.2 (-0.2-0.6) |  |
| **Study 2** |  |  |  |  |  |  |  |  |
| Primary infections in unvaccinated individuals | 46 | 20.9 |  | 17.3-25.6 | 21.7 (20.0-23.3) | 5.5 |  | <0.001 |
| mRNA-1273-vaccine breakthrough infections | 46 | 27.5 | 6.6 | 19.9-32.1 | 26.6 (24.6-28.6) | 6.7 | 4.9 (2.4-7.4) |  |
| **Study 3** |  |  |  |  |  |  |  |  |
| BNT162b2-vaccine breakthrough infections | 39 | 19.4 |  | 17.1-24.0 | 21.3 (19.3-23.2) | 6.0 |  | <0.001 |
| mRNA-1273-vaccine breakthrough infections | 39 | 27.5 | 8.1 | 21.8-32.0 | 26.6 (24.5-28.7) | 6.5 | 5.3 (2.5-8.2) |  |
| **Study 4** |  |  |  |  |  |  |  |  |
| Primary infections in unvaccinated individuals | 364 | 22.1 |  | 17.5-27.4 | 22.7 (22.1-23.3) | 5.9 |  | <0.001 |
| Reinfections in unvaccinated individuals | 364 | 27.2 | 5.1 | 20.3-32.9 | 26.5 (25.8-27.2) | 6.8 | 3.8 (2.8-4.7) |  |
| **Study 5** |  |  |  |  |  |  |  |  |
| BNT162b2-vaccine breakthrough infections | 204 | 22.9 |  | 17.8-29.3 | 23.7 (22.9-24.6) | 6.1 |  |  |
| Reinfections in unvaccinated individuals | 204 | 25.3 | 2.4 | 19.4-31.6 | 25.6 (24.6-26.5) | 6.6 | 1.8 (0.6-3.1) | 0.004 |
| **Study 6** |  |  |  |  |  |  |  |  |
| mRNA-1273-vaccine breakthrough infections | 13 | 21.9 |  | 18.3-28.5 | 23.5 (19.6-27.4) | 6.4 |  | 0.016 |
| Reinfections in unvaccinated individuals | 13 | 33.1 | 11.2 | 27.1-34.3 | 30.0 (26.2-33.7) | 6.3 | 6.4 (1.3-11.6) |  |

Abbreviations: Ct, cycle threshold; IQR, interquartile range; RT-qPCR, real time reverse transcription polymerase chain reaction; SD, standard deviation.
\*Study groups were matched in a 1:1 ratio by sex, 10-year age group, reason for RT-qPCR testing, and RT-qPCR test calendar week.

### Differences in SARS-CoV-2 viral load and infectiousness

These differences in Ct values can be translated into approximate *relative* differences in viral load (Methods). Since the mathematical formula linking viral load to Ct value has an exponential form (viral load is proportional to 2^−*n*^ where *n* is the RT-qPCR Ct value)^21^, small differences in Ct value represent large differences in viral load.

Empirically, SARS-CoV-2 viral load strongly correlates with culturable virus^10^, just as viral load for other viruses, such as HIV, is a measure of its infectiousness^22–24^. Therefore, relative differences in viral load can be considered as a proxy measure of relative differences in infectiousness (Methods), that is *VE*_*I*_ ^7^. For instance, assuming that SARS-CoV-2 infectiousness is linearly proportional to viral load, the simplest default assumption, a Ct value higher by 1.0 cycle corresponds to a lower viral load (and infectiousness) by a factor of 2 (2^−1.0^ = 1/2), that is a *VE*_*I*_ = 50%.

Table 5 shows approximate estimates for the relative infectiousness of SARS-CoV-2 infections across the cohorts of primary infections, reinfections, BNT162b2 breakthrough infections, and mRNA-1273 breakthrough infections, assuming that SARS-CoV-2 infectiousness is linearly proportional to viral load. Meanwhile, Table 6 shows the same estimates in a sensitivity analysis assuming that infectiousness is (non-linearly) proportional to some power of viral load (*vl*^*α*^), just as for HIV infection which has perhaps the best characterized relationship between viral load and infectiousness in the literature^22–24^. Here, the exponent *α* has been estimated at around 0.4^22- 24^—that is infectiousness is approximately proportional to the square rate of viral load rather than the viral load itself.

In both sets of estimates, breakthrough infections in those vaccinated or who had a prior infection had substantially lower infectiousness than primary infections in unvaccinated persons. For example, in the randomly diagnosed infections and assuming that infectiousness is linearly proportional to viral load, BNT162b2 breakthrough infections, mRNA-1273 breakthrough infections, and reinfections were only 0.41-fold (95% CI: 0.29-0.54), 0.11-fold (95% CI: 0.04-0.29), and 0.06-fold (95% CI: 0.04-0.09) as infectious as primary infections, respectively. The different comparisons suggest an overall hierarchy, present for both asymptomatic and symptomatic infections, where primary infections in unvaccinated persons are most infectious, followed by BNT162b2 breakthrough infections, mRNA-1273 breakthrough infections, and finally reinfections in unvaccinated persons.

## Discussion

Breakthrough infections in those vaccinated or who had a prior infection have higher RT-qPCR Ct values than primary infections in unvaccinated persons. While these breakthrough infections are not uncommon, the results indicate that they have lower viral load and are less likely to be infectious than primary infections. While some of these breakthrough infections could lead to secondary transmissions, and indeed some of them did have high viral loads (Figure 3 and Supplementary Figure 1 and 2), the risk of onward transmission appears to be substantially reduced, compared to primary infections. Thus, they are of less public health concern.

A consequence of these findings is that the public health benefits of vaccination may be underestimated. In addition to the conventional vaccine efficacy against acquisition of infection (*VE*_*S*_) that is assessed in randomized clinical trials^1–3^, there is an additional “breakthrough” efficacy against transmission (*VE*_*I*_) that augments the benefits of *VE*_*S*_ (Tables 5 and 6), at least for the BNT162b2 and mRNA-1273 vaccines investigated in this study. The existence of this additional *VE*_*I*_ efficacy implies that the reproduction number (*R*_0_) after vaccination is lower than current estimates^7^. SARS-CoV-2 incidence may decline faster with vaccine scale up^7^, and herd immunity may be easier to achieve, at somewhat lower vaccine coverage than previously predicted^7^.

One finding of this study is that there appears to be a hierarchy in infectiousness of SARS-CoV-2 infections, where primary infections in unvaccinated persons are most infectious, followed by BNT162b2 breakthrough infections, mRNA-1273 breakthrough infections, and finally reinfections in unvaccinated persons. Strikingly, this hierarchy is the mirror image of the hierarchy observed in the efficacy against acquisition of infection. In our earlier studies on these national cohorts in Qatar, we found that those vaccinated with BNT162b2 had (relatively) the lowest protection against acquisition of infection (at 75% against the B.1.351 (Beta) variant^4,11^ that dominated incidence since the onset of vaccination^19,20^), while protection against acquisition of infection was considerably higher among those vaccinated with mRNA-1273^5^ and those with a prior infection^13–15,25^. This may indicate that both *VE*_*S*_ and *VE*_*I*_ are essentially inter-related manifestations of the strength of the vaccine-induced (or natural-infection-induced) immune response. When the immune response is strong against acquisition of infection, it also appears strong at reducing viral replication upon acquisition of the virus, leading to lower viral load and faster infection clearance. Thus, less secondary transmission occurs in breakthrough infections.

This study has limitations. The number of documented mRNA-1273 breakthrough infections was small, thereby limiting the statistical precision of the comparisons involving these infections, leading to estimates with wider 95% confidence intervals, and perhaps making them more prone to bias. The study was implemented on documented infections, but other infections may have occurred and gone undocumented. It is possible that breakthrough infections in those vaccinated or who had a prior infection are less likely to be documented, perhaps because of minimal/mild or no symptoms. However, with the high rate of PCR testing in Qatar, the majority of infections are identified not because of testing symptomatic cases, but because of testing for other reasons, such as random testing campaigns, contact tracing, individual requests, routine healthcare testing, pre-travel, and at ports of entry. The results of our study were also consistent when we included only the randomly diagnosed infections.

Imperfect assay sensitivity and specificity of RT-qPCR testing may have affected infection ascertainment, but RT-qPCR testing was performed using a validated commercial platform that has been used globally and has essentially 100% sensitivity and specificity^26^ (Methods). While it is established that SARS-CoV-2 viral load strongly correlates with culturable virus^10^ and infectiousness, the exact mathematical relationship linking viral load to infectiousness is not known. Therefore, we generated estimates for the relative infectiousness assuming a linear dependence on viral load, and alternatively assuming a non-linear (power-law) dependence on viral load, similar to that rigorously delineated for HIV infection^22–24^. Estimates for the relative infectiousness presented in Tables 4 and 5 are thus approximate and qualitative in nature, and should not be seen as precise quantitative measures.

**Table 4.** Relative infectiousness of all RT-qPCR confirmed infections, only the randomly diagnosed (asymptomatic) infections, and only the symptomatic infections across the six pairwise comparisons between primary infections in unvaccinated individuals, reinfections in unvaccinated individuals, BNT162b2-vaccine breakthrough infections, and mRNA-1273-vaccine breakthrough infections. These estimates assume that SARS-CoV-2 infectiousness is linearly proportional to viral load. Estimates in the sensitivity analysis assuming that infectiousness is (non-linearly) proportional to some power of viral load, similar to that for HIV^22–24^, are found in Table 5.

| Pairwise comparison | Relative infectiousness (95% CI) |  |  |
| --- | --- | --- | --- |
|  | All RT-qPCR confirmed infections | Randomly diagnosed (asymptomatic) infection | Symptomatic infections |
| Infectiousness of BNT162b2-vaccine breakthrough infections relative to primary infections in unvaccinated individuals | 0.50 (0.44-0.62) | 0.41 (0.29-0.54) | 0.87 (0.66-1.15) |
| Infectiousness of mRNA-1273-vaccine breakthrough infections relative to primary infections in unvaccinated individuals | 0.09 (0.04-0.19) | 0.11 (0.04-0.29) | 0.03 (0.01-0.19) |
| Infectiousness of mRNA-1273-vaccine breakthrough infections relative to BNT162b2-vaccine breakthrough infections | 0.12 (0.05-0.27) | 0.20 (0.08-0.57) | 0.03 (0.00-0.18) |
| Infectiousness of reinfections in unvaccinated individuals relative to primary infections in unvaccinated individuals | 0.08 (0.05-0.09) | 0.06 (0.04-0.09) | 0.07 (0.04-0.1) |
| Infectiousness of BNT162b2-vaccine breakthrough infections relative to reinfections in unvaccinated individuals | 0.31 (0.19-0.47) | 0.25 (0.14-0.47) | 0.29 (0.12-0.66) |
| Infectiousness of mRNA-1273-vaccine breakthrough infections relative to reinfections in unvaccinated individuals | 0.25 (0.07-0.87) | 0.31 (0.07-1.32) | 0.01 (0.00-0.41) |

Abbreviations: RT-qPCR, real time reverse transcription polymerase chain reaction

**Table 5.** Sensitivity analysis for estimates of relative infectiousness. Relative infectiousness of all RT-qPCR confirmed infections, only the randomly diagnosed (asymptomatic) infections, and only the symptomatic infections across the six pairwise comparisons between primary infections in unvaccinated individuals, reinfections in unvaccinated individuals, BNT162b2-vaccine breakthrough infections, and mRNA-1273-vaccine breakthrough infections. The sensitivity analysis estimates assume that SARS-CoV-2 infectiousness is (non-linearly) proportional to some power of viral load (*vl*^*α*^), just as for HIV, where the exponent *α* has been estimated at around 0.4^22–24^. Estimates assuming that SARS-CoV-2 infectiousness is linearly proportional to viral load (that is *α* =1) are found in Table 4.

| Pairwise comparison | Relative infectiousness (95% CI) |  |  |
| --- | --- | --- | --- |
|  | All RT-qPCR confirmed infections | Randomly diagnosed (asymptomatic) infection | Symptomatic infections |
| Infectiousness of BNT162b2-vaccine breakthrough infections relative to primary infections in unvaccinated individuals | 0.76 (0.72-0.82) | 0.70 (0.61-0.78) | 0.95 (0.85-1.06) |
| Infectiousness of mRNA-1273-vaccine breakthrough infections relative to primary infections in unvaccinated individuals | 0.38 (0.28-0.51) | 0.41 (0.29-0.61) | 0.26 (0.13-0.51) |
| Infectiousness of mRNA-1273-vaccine breakthrough infections relative to BNT162b2-vaccine breakthrough infections | 0.42 (0.30-0.59) | 0.53 (0.36-0.80) | 0.23 (0.10-0.50) |
| Infectiousness of reinfections in unvaccinated individuals relative to primary infections in unvaccinated individuals | 0.36 (0.31-0.39) | 0.33 (0.28-0.39) | 0.35 (0.27-0.46) |
| Infectiousness of BNT162b2-vaccine breakthrough infections relative to reinfections in unvaccinated individuals | 0.62 (0.51-0.74) | 0.57 (0.46-0.74) | 0.61 (0.42-0.85) |
| Infectiousness of mRNA-1273-vaccine breakthrough infections relative to reinfections in unvaccinated individuals | 0.57 (0.34-0.95) | 0.62 (0.35-1.12) | 0.17 (0.04-0.70) |

Abbreviations: RT-qPCR, real time reverse transcription polymerase chain reaction

Unlike blinded, randomized clinical trials, the investigated observational cohorts were neither blinded nor randomized. Our cohorts predominantly included working-age adults; therefore, results may not necessarily be generalizable to other population groups, such as children or the elderly. Matching was done for sex, age, reason for the RT-qPCR testing, and calendar week of the RT-qPCR test, but could not be done for other factors, such as comorbidities, as these were not available to study investigators. But inclusion of additional factors in the matching would have considerably reduced the sample sizes—breakthrough infections in those vaccinated or who had a prior infection are relatively uncommon. It is noteworthy that matching by age and sex may have served as a proxy for matching by co-morbidity, as co-morbidities are associated with older age and may differ between women and men.

In conclusion, prior immunity, whether due to vaccination or prior infection, is associated with lower SARS-CoV-2 viral load upon infection. While breakthrough infections have been observed globally, they appear less infectious than primary infections; thus, they constitute a lesser public health concern. The public health benefits of vaccination are underestimated, as both the BNT162b2 and mRNA-1273 vaccines seem not only to protect against acquisition of infection, but also against transmission of infection. These findings justify optimism and stress the urgency to scale up vaccination globally in order to robustly control infection transmission and the extent of the pandemic.

## Data Availability

The dataset of this study is a property of the Qatar Ministry of Public Health that was provided to the researchers through a restricted-access agreement that prevents sharing the dataset with a third party or publicly. Future access to this dataset can be considered through a direct application for data access to Her Excellency the Minister of Public Health (https://www.moph.gov.qa/english/Pages/default.aspx). Aggregate data are available within the manuscript and its Supplementary information.

## Acknowledgements

We acknowledge the many dedicated individuals at Hamad Medical Corporation, the Ministry of Public Health, the Primary Health Care Corporation, and the Qatar Biobank for their diligent efforts and contributions to make this study possible. The authors are grateful for support from the Biomedical Research Program and the Biostatistics, Epidemiology, and Biomathematics Research Core, both at Weill Cornell Medicine-Qatar, as well as for support provided by the Ministry of Public Health and Hamad Medical Corporation. The authors are also grateful for the Qatar Genome Programme for supporting the viral genome sequencing. The funders of the study had no role in study design, data collection, data analysis, data interpretation, or writing of the article. Statements made herein are solely the responsibility of the authors.

## Author contributions

LJA conceived and co-designed the study, led the statistical analyses, and co-wrote the first draft of the article. HC co-designed the study, performed the statistical analyses, and co-wrote the first draft of the article. All authors contributed to data collection and acquisition, database development, discussion and interpretation of the results, and to the writing of the manuscript. All authors have read and approved the final manuscript.

## Methods

### Data sources and study design

Analyses were conducted using the centralized, integrated, and standardized national severe acute respiratory syndrome coronavirus 2 (SARS-CoV-2) databases compiled at Hamad Medical Corporation (HMC), the main public healthcare provider and the nationally designated provider for all Coronavirus Disease 2019 (COVID-19) health care needs. Through a nationwide digital health information platform, these databases are complete, and have captured all SARS-CoV-2-related data as well as related demographic details with no missing information since the start of the epidemic, including all records of polymerase chain reaction (PCR) testing, antibody testing, COVID-19 hospitalizations, vaccinations, infection severity classification per World Health Organization (WHO) guidelines^27^ (performed by trained medical personnel through individual chart reviews), and COVID-19 deaths, also assessed per WHO guidelines^28^.

Every real time reverse transcription-PCR (RT-qPCR) test conducted in Qatar, regardless of location (outpatient clinic, drive-thru, or hospital, etc.), is classified on the basis of symptoms and the reason for testing (clinical symptoms, contact tracing, random testing campaigns (surveys), individual requests, routine healthcare testing, pre-travel, and port of entry). Qatar has unique demographics by sex and nationality, since expatriates from over 150 countries comprise 89% of the population^17,29^.

The BNT162b2 and mRNA-1273 vaccines have been the vaccines of choice in the national immunization campaign in Qatar^4,5,11,12^. Nearly all individuals in the cohorts of this study were vaccinated (free of charge) in Qatar, rather than elsewhere. In rare situations where an individual received vaccination outside Qatar, that individual’s vaccination details were still recorded in the health system at the port of entry upon return to Qatar, given the national requirements and to benefit from privileges associated with vaccination, such as quarantine exemption^12^.

Leveraging the national databases, effects of vaccination and of prior infection on SARS-CoV-2 infectiousness were investigated by comparing the RT-qPCR cycle threshold (Ct) values in matched cohorts of primary infections in unvaccinated individuals, reinfections in unvaccinated individuals, BNT162b2 breakthrough infections, and mRNA-1273 breakthrough infections. These types of infection are defined in the main text.

In total, six types of pairwise comparisons were conducted on the cohorts of this study, after matching in a 1:1 ratio by sex, 10-year age group, reason for RT-qPCR testing, and calendar week of the RT-qPCR test, to control for differences in biology by sex and age, as well as exposure risk^17,18^ and variant exposure^4,19,20^. It is noteworthy that the first SARS-CoV-2 epidemic wave in Qatar occurred before introduction of any variant of concern and peaked in late May, 2020^17,18^. The second wave was triggered by introduction and expansion of the B.1.1.7 (Alpha^30^) variant and peaked in early March, 2021^4,5,19,20^. The third wave was dominated by the B.1.351 (Beta^30^) variant, and peaked in the first week of April, 2021^4,5,19,20^. The B.1.617.2 (Delta^30^) variant has been introduced only recently in Qatar, and as of July 11, 2021, it remains at low incidence^19,20^. There is no evidence that any other variant of concern is or has been responsible for appreciable community transmission in Qatar^19,20^.

Comparisons across the cohorts of infection were implemented for all RT-qPCR-confirmed infections, for only the symptomatic infections defined as RT-qPCR-positive tests conducted because of clinical suspicion due to symptoms compatible with a respiratory tract infection, and for only the *randomly diagnosed* (asymptomatic) infections defined as RT-qPCR-positive tests conducted with no prior reason to suspect infection and no reported symptoms compatible with a respiratory tract infection. The latter was strictly defined as an RT-qPCR-positive test conducted as part of a survey (random testing campaigns), for routine health care testing, as a pre-travel requirement, or at a port of entry upon arrival in the country^4,5,12,17^.

All records of RT-qPCR testing in Qatar were examined in this study, but only samples of matched cohorts were included in the analysis. Individuals with a record of a SARS-CoV-2 antibody-positive test before the first RT-qPCR-positive test were excluded from analysis of those with primary infections. Individuals with a record of vaccination before the reinfection diagnosis were excluded from the analysis of those with reinfection. Only breakthrough infections in fully vaccinated individuals were included in the analysis. Being fully vaccinated was defined as completion of ≥14 days after the second dose. Only individuals who had their first and second doses with the same vaccine were included in the analysis.

Further background on Qatar’s epidemic, such as on reinfections^13,14^, national seroprevalence surveys^17,31–33^, RT-qPCR surveys^17^, and other epidemiological studies can be found in previous publications on this epidemic^4,5,11,17,18,34–40^.

Reporting of the study followed the STROBE guidelines (Supplementary Table 3).

### Laboratory methods

Nasopharyngeal and/or oropharyngeal swabs (Huachenyang Technology, China) were collected for PCR testing and placed in Universal Transport Medium (UTM). Aliquots of UTM were: extracted on a QIAsymphony platform (QIAGEN, USA) and tested with real-time reverse-transcription PCR (RT-qPCR) using TaqPath™ COVID-19 Combo Kits (100% sensitivity and specificity^26^; Thermo Fisher Scientific, USA) on an ABI 7500 FAST (ThermoFisher, USA); extracted using a custom protocol^41^ on a Hamilton Microlab STAR (Hamilton, USA) and tested using AccuPower SARS-CoV-2 Real-Time RT-PCR Kits (100% sensitivity and specificity^42^; Bioneer, Korea) on an ABI 7500 FAST; or loaded directly into a Roche cobas® 6800 system and assayed with a cobas® SARS-CoV-2 Test (95% sensitivity, 100% specificity^43^; Roche, Switzerland). The first assay targets the viral S, N, and ORF1ab regions. The second targets the viral RdRp and E-gene regions, and the third targets the ORF1ab and E-gene regions.

All tests were conducted at the HMC Central Laboratory or Sidra Medicine Laboratory, following standardized protocols.

Only the RT-qPCR-confirmed infections diagnosed using the TaqPath COVID-19 Combo Kits platform (Thermo Fisher Scientific, USA^26^) were included in the analysis, for standardization of Ct values. This platform processes >85% of all RT-qPCR tests in Qatar and reports individual Ct values for each of the N, ORF1ab, and S genes^4,5,26^. The average of these three Ct values was included in the analysis. The correlation between each pair of these Ct values across all RT-qPCR-positive tests was very strong with a Pearson correlation coefficient ≥0.976. In the case of a gene “target failure”, and specifically an S-gene “target failure” that was the defining characteristic of the B.1.1.7 cases^44–46^, the average determined using only the two Ct values of the N and ORF1ab genes.

### Statistical analysis

Socio-demographic characteristics of study samples were described using frequency distributions and measures of central tendency. Differences in proportions across categorical variables between study groups were evaluated using Chi-square tests. The distributions of the RT-qPCR Ct values were illustrated using scatter plots and boxplots, and summarized using measures of central tendency and dispersion. Mean differences in the RT-qPCR Ct values between study groups and associated 95% confidence intervals (CIs) were calculated using independent t-tests. Two-sided p-value of <0.05 indicated a significant association. Statistical analyses were conducted in STATA/SE version 17.0^47^.

### RT-qPCR Ct value, viral load, and SARS-CoV-2 infectiousness

SARS-CoV-2 viral load (*vl*) can be approximately estimated from the RT-qPCR Ct value (*n*) using the mathematical formula^21,26^:

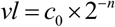

where *c*_0_ is a constant that depends on each RT-qPCR platform and is experimentally derived factoring specimen buffers and extraction method among other factors.

Accordingly, in a pairwise comparison of RT-qPCR-confirmed infections in two cohorts, the ratio of the two viral loads is independent of the constant *c*_0_ and is given by

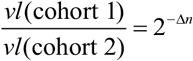

where Δ*n* is the difference in the RT-qPCR Ct values between these two cohorts.

SARS-CoV-2 viral load correlates strongly with cultivable virus^10^, and thus infectiousness, just as viral load for other viruses, such as HIV, is a measure of its infectiousness^22–24^. Therefore, the ratio of two viral loads in two cohorts can be considered to provide a measure of the relative infectiousness of these two cohorts. This implicitly assumes that SARS-CoV-2 infectiousness is linearly proportional to *vl*, the simplest default assumption for lack of data. This assumption was used in the baseline analysis for the relative infectiousness.

However, infectiousness could be related to viral load through a non-linear mathematical function. For HIV infection for instance, which has perhaps the best characterized relationship between viral load and infectiousness in the literature^22–24^, infectiousness is (non-linearly) proportional to *vl*^*α*^ where the exponent *α* has been estimated at 0.3-0.5^22–24^—that is infectiousness is approximately proportional to the square rate of viral load rather than the viral load itself. In a sensitivity analysis, we estimated SARS-CoV-2 relative infectiousness assuming that infectiousness is proportional to *vl*^*α*^ with an *α* = 0.4, similar to that for HIV^22–24^, to provide an alternative estimate assuming a power-law mathematical function linking viral load to infectiousness.

## Ethical approvals

The study was approved by the Hamad Medical Corporation and Weill Cornell Medicine-Qatar Institutional Review Boards with waiver of informed consent.

## Supplementary Material

**Supplementary Table 1.**
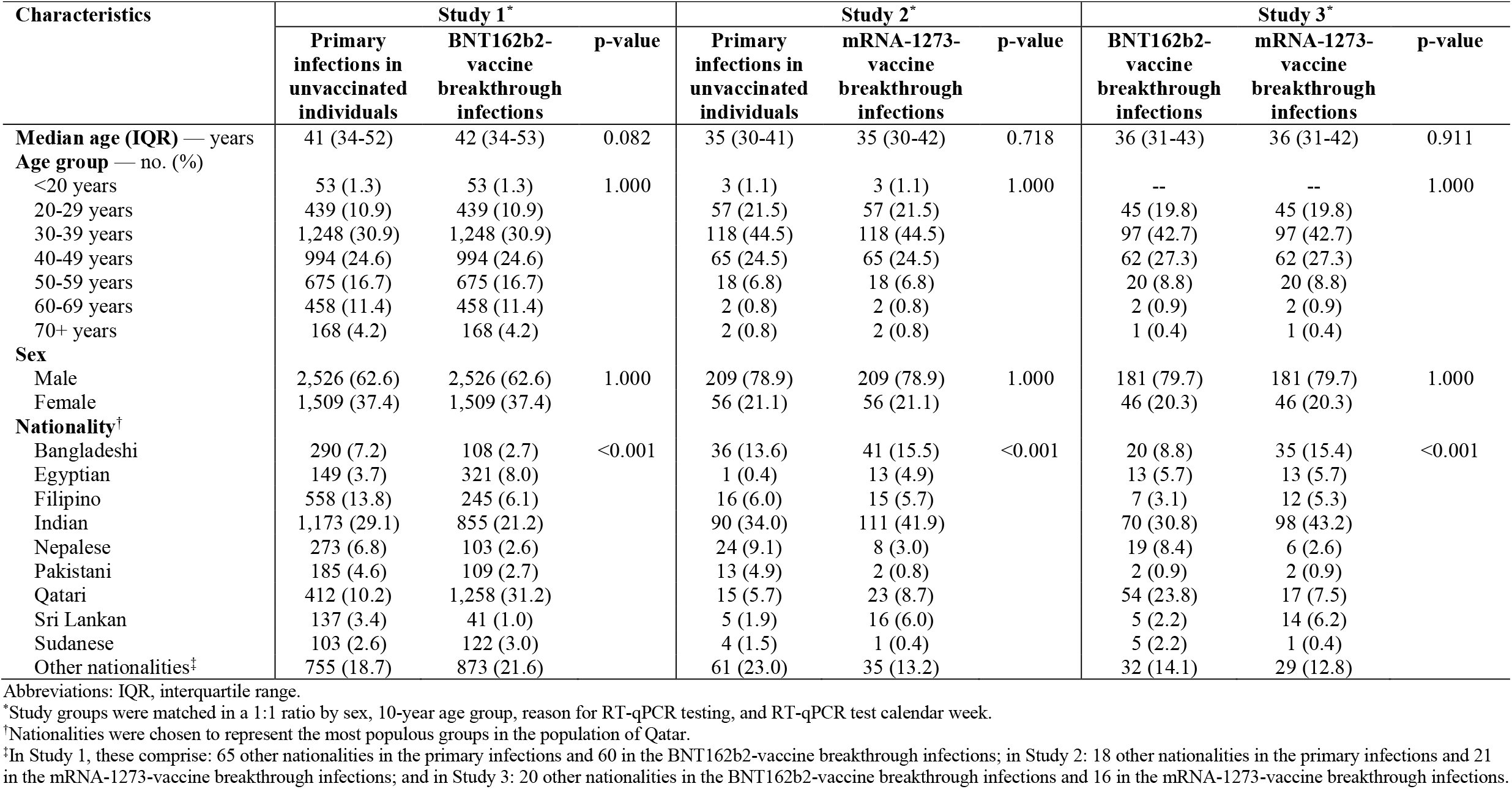
Demographic characteristics of the study populations in the three pairwise comparisons between primary infections in unvaccinated individuals, BNT162b2-vaccine breakthrough infections, and mRNA-1273-vaccine breakthrough infections.

| Characteristics | Study 1* |  |  | Study 2* |  |  | Study 3* |  |  |
| --- | --- | --- | --- | --- | --- | --- | --- | --- | --- |
|  | Primary infections in unvaccinated individuals | BNT162b2-vaccine breakthrough infections | p-value | Primary infections in unvaccinated individuals | mRNA-1273-vaccine breakthrough infections | p-value | BNT162b2-vaccine breakthrough infections | mRNA-1273-vaccine breakthrough infections | p-value |
| **Median age (IQR) — years** | 41 (34-52) | 42 (34-53) | 0.082 | 35 (30-41) | 35 (30-42) | 0.718 | 36 (31-43) | 36 (31-42) | 0.911 |
| **Age group — no. (%)** |  |  |  |  |  |  |  |  |  |
| <20 years | 53 (1.3) | 53 (1.3) | 1.000 | 3 (1.1) | 3 (1.1) | 1.000 | -- | -- | 1.000 |
| 20-29 years | 439 (10.9) | 439 (10.9) |  | 57 (21.5) | 57 (21.5) |  | 45 (19.8) | 45 (19.8) |  |
| 30-39 years | 1,248 (30.9) | 1,248 (30.9) |  | 118 (44.5) | 118 (44.5) |  | 97 (42.7) | 97 (42.7) |  |
| 40-49 years | 994 (24.6) | 994 (24.6) |  | 65 (24.5) | 65 (24.5) |  | 62 (27.3) | 62 (27.3) |  |
| 50-59 years | 675 (16.7) | 675 (16.7) |  | 18 (6.8) | 18 (6.8) |  | 20 (8.8) | 20 (8.8) |  |
| 60-69 years | 458 (11.4) | 458 (11.4) |  | 2 (0.8) | 2 (0.8) |  | 2 (0.9) | 2 (0.9) |  |
| 70+ years | 168 (4.2) | 168 (4.2) |  | 2 (0.8) | 2 (0.8) |  | 1 (0.4) | 1 (0.4) |  |
| **Sex** |  |  |  |  |  |  |  |  |  |
| Male | 2,526 (62.6) | 2,526 (62.6) | 1.000 | 209 (78.9) | 209 (78.9) | 1.000 | 181 (79.7) | 181 (79.7) | 1.000 |
| Female | 1,509 (37.4) | 1,509 (37.4) |  | 56 (21.1) | 56 (21.1) |  | 46 (20.3) | 46 (20.3) |  |
| **Nationality<sup>†</sup>** |  |  |  |  |  |  |  |  |  |
| Bangladeshi | 290 (7.2) | 108 (2.7) | <0.001 | 36 (13.6) | 41 (15.5) | <0.001 | 20 (8.8) | 35 (15.4) | <0.001 |
| Egyptian | 149 (3.7) | 321 (8.0) |  | 1 (0.4) | 13 (4.9) |  | 13 (5.7) | 13 (5.7) |  |
| Filipino | 558 (13.8) | 245 (6.1) |  | 16 (6.0) | 15 (5.7) |  | 7 (3.1) | 12 (5.3) |  |
| Indian | 1,173 (29.1) | 855 (21.2) |  | 90 (34.0) | 111 (41.9) |  | 70 (30.8) | 98 (43.2) |  |
| Nepalese | 273 (6.8) | 103 (2.6) |  | 24 (9.1) | 8 (3.0) |  | 19 (8.4) | 6 (2.6) |  |
| Pakistani | 185 (4.6) | 109 (2.7) |  | 13 (4.9) | 2 (0.8) |  | 2 (0.9) | 2 (0.9) |  |
| Qatari | 412 (10.2) | 1,258 (31.2) |  | 15 (5.7) | 23 (8.7) |  | 54 (23.8) | 17 (7.5) |  |
| Sri Lankan | 137 (3.4) | 41 (1.0) |  | 5 (1.9) | 16 (6.0) |  | 5 (2.2) | 14 (6.2) |  |
| Sudanese | 103 (2.6) | 122 (3.0) |  | 4 (1.5) | 1 (0.4) |  | 5 (2.2) | 1 (0.4) |  |
| Other nationalities<sup>‡</sup> | 755 (18.7) | 873 (21.6) |  | 61 (23.0) | 35 (13.2) |  | 32 (14.1) | 29 (12.8) |  |

Abbreviations: IQR, interquartile range.
\*Study groups were matched in a 1:1 ratio by sex, 10-year age group, reason for RT-qPCR testing, and RT-qPCR test calendar week.
<sup>†</sup>Nationalities were chosen to represent the most populous groups in the population of Qatar.
<sup>‡</sup>In Study 1, these comprise: 65 other nationalities in the primary infections and 60 in the BNT162b2-vaccine breakthrough infections; in Study 2: 18 other nationalities in the primary infections and 21 in the mRNA-1273-vaccine breakthrough infections; and in Study 3: 20 other nationalities in the BNT162b2-vaccine breakthrough infections and 16 in the mRNA-1273-vaccine breakthrough infections.

**Supplementary Table 2.**
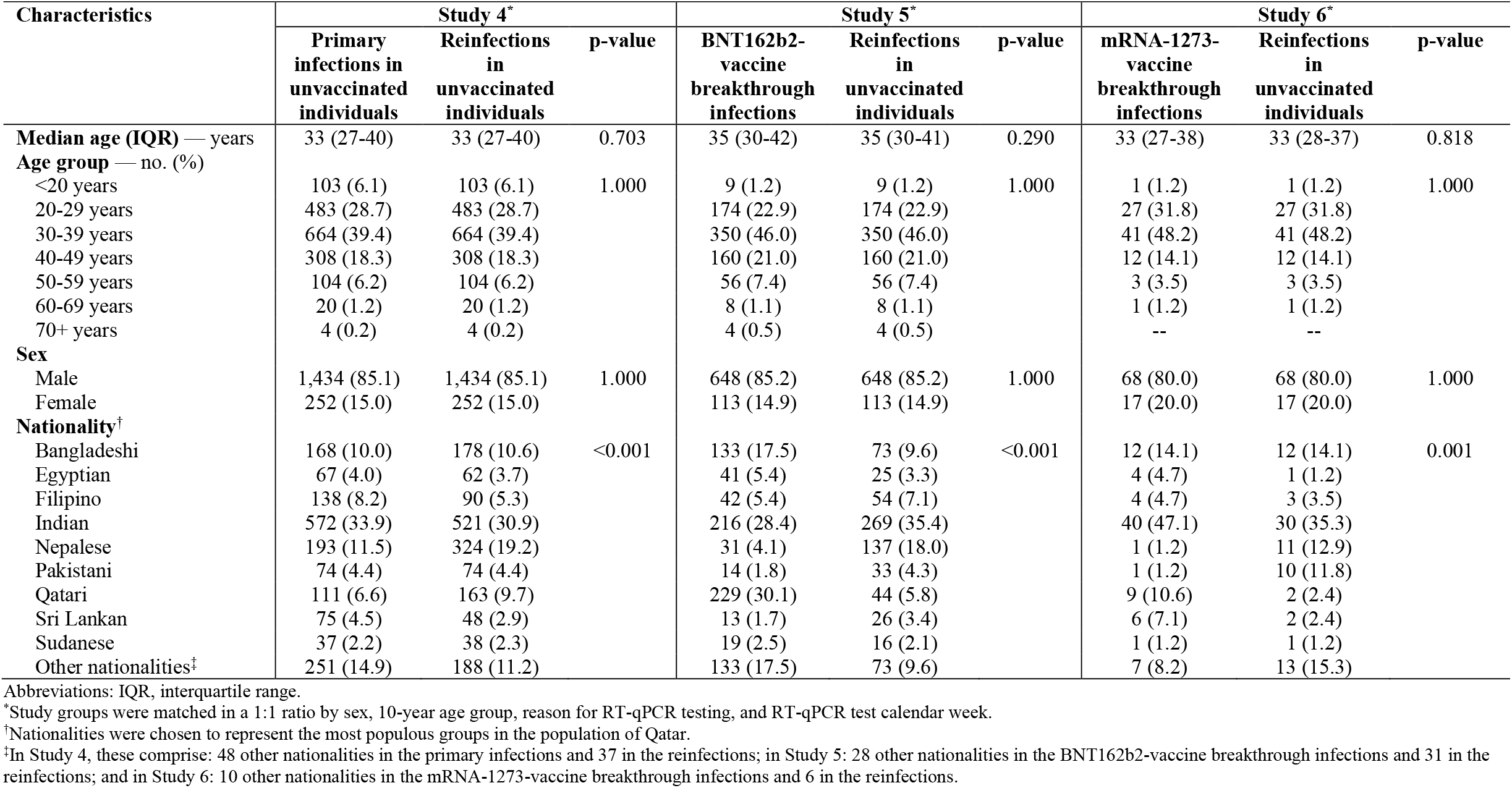
Demographic characteristics of the study populations in three pairwise comparisons between primary infections in unvaccinated individuals, reinfections in unvaccinated individuals, BNT162b2-vaccine breakthrough infections, and mRNA-1273-vaccine breakthrough infections.

**Supplementary Figure 1.**
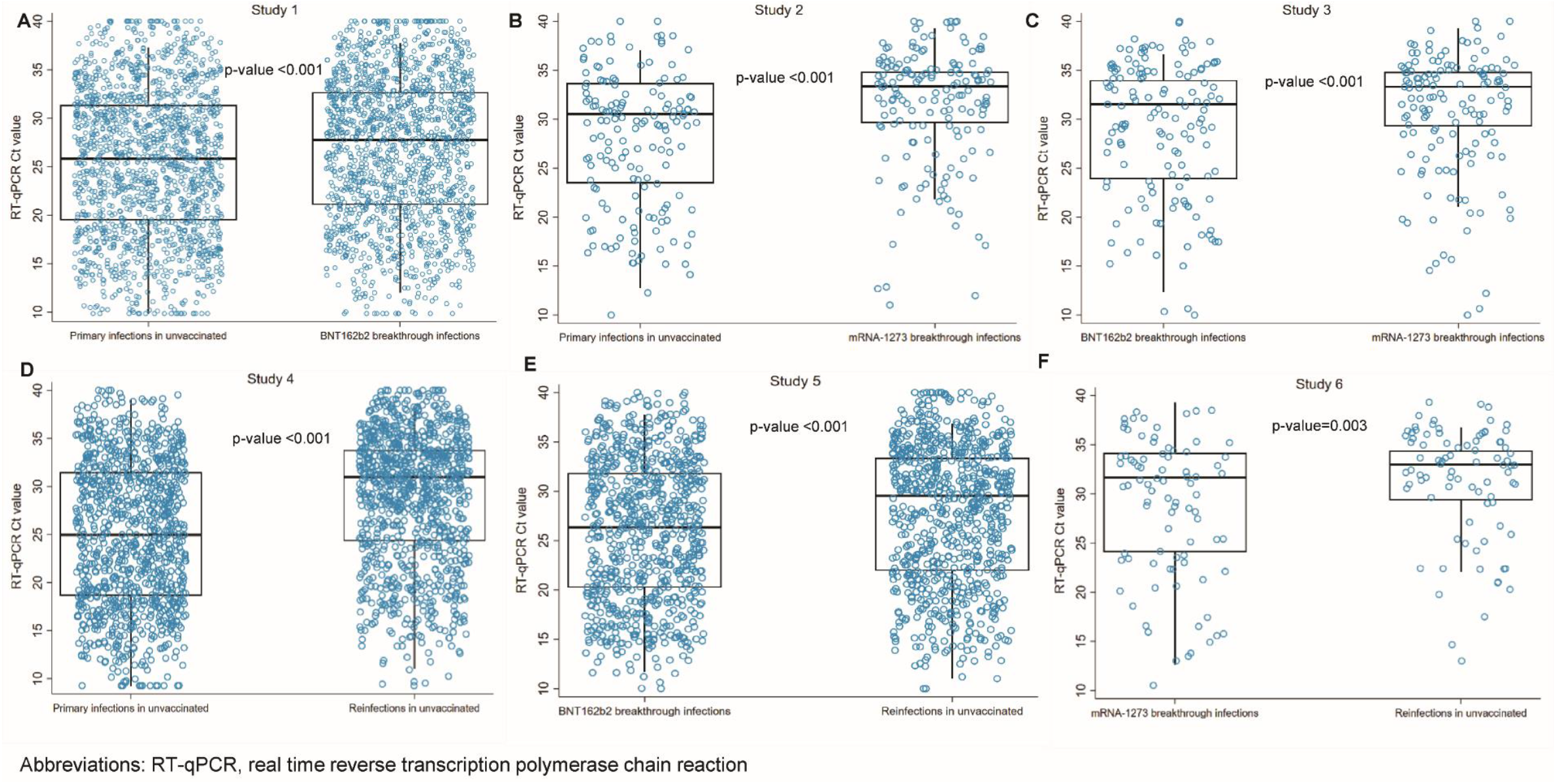
RT-qPCR Ct values in all confirmed infections, regardless of the reason for the RT-qPCR testing. Distribution of these Ct values in the six pairwise comparisons between primary infections in unvaccinated individuals, reinfections in unvaccinated individuals, BNT162b2-vaccine breakthrough infections, and mRNA-1273-vaccine breakthrough infections. Boxplots center lines indicate the median Ct values, box limits indicate the 25% and 75% quartiles, and whiskers indicate maximum and minimum observations within 1.5 of interquartile range.

**Supplementary Figure 2.**
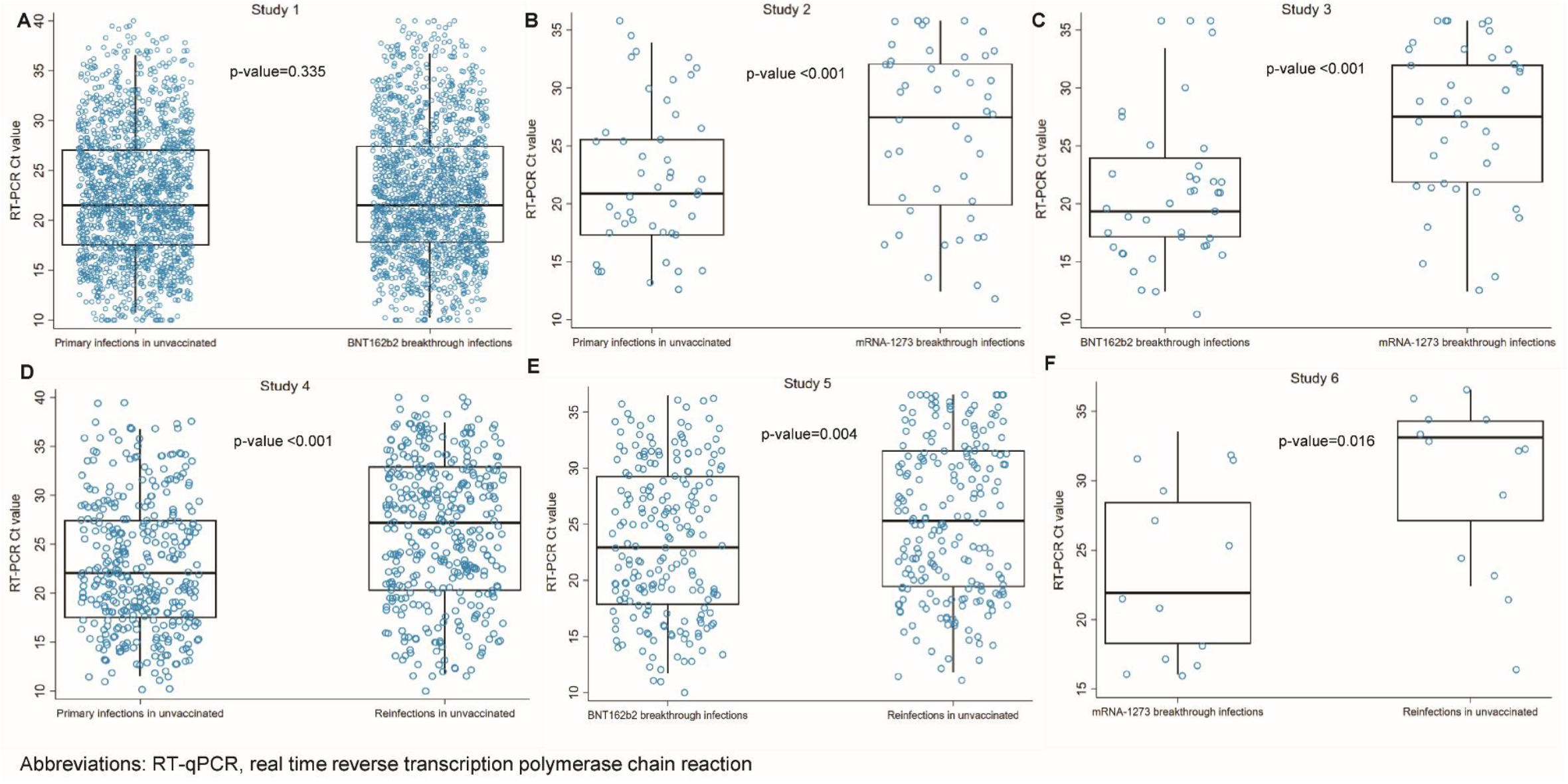
RT-qPCR Ct values in the symptomatic SARS-CoV-2 infections. Distribution of these Ct values in the six pairwise comparisons between primary infections in unvaccinated individuals, reinfections in unvaccinated individuals, BNT162b2-vaccine breakthrough infections, and mRNA-1273-vaccine breakthrough infections. A symptomatic infection was defined as an RT-qPCR-positive test conducted because of clinical suspicion due to presence of symptoms compatible with a respiratory tract infection. Boxplots center lines indicate the median Ct values, box limits indicate the 25% and 75% quartiles, and whiskers indicate maximum and minimum observations within 1.5 of interquartile range.

**Supplementary Table 3.** STROBE checklist.

|  | Item No | Recommendation | Main Text Page # |
| --- | --- | --- | --- |
| Title and abstract | 1 | (a) Indicate the study’s design with a commonly used term in the title or the abstract | 2 |
|  |  | (b) Provide in the abstract an informative and balanced summary of what was done and what was found | 2 |
| Introduction |  |  |  |
| Background/rationale | 2 | Explain the scientific background and rationale for the investigation being reported | 3 |
| Objectives | 3 | State specific objectives, including any prespecified hypotheses | 3-4 |
| Methods |  |  |  |
| Study design | 4 | Present key elements of study design early in the paper | 3-4, 23-24 |
| Setting | 5 | Describe the setting, locations, and relevant dates, including periods of recruitment, exposure, follow-up, and data collection | 3-4, 23-24 |
| Participants | 6 | (a) Give the eligibility criteria, and the sources and methods of selection of participants | 23-24 |
| Variables | 7 | Clearly define all outcomes, exposures, predictors, potential confounders, and effect modifiers. Give diagnostic criteria, if applicable | 23-27 |
| Data sources/<br>measurement | 8* | For each variable of interest, give sources of data and details of methods of assessment (measurement). Describe comparability of assessment methods if there is more than one group | 23-25 & Supp. Tables 1 & 2 |
| Bias | 9 | Describe any efforts to address potential sources of bias | 24-27 |
| Study size | 10 | Explain how the study size was arrived at | 23-25 & Figures 1-2 |
| Quantitative variables | 11 | Explain how quantitative variables were handled in the analyses. If applicable, describe which groupings were chosen and why | 26-27 & Supp. Tables 1 & 2 |
| Statistical methods | 12 | (a) Describe all statistical methods, including those used to control for confounding | 24-25 & 26-27 |
|  |  | (b) Describe any methods used to examine subgroups and interactions | 24-25 |
|  |  | (c) Explain how missing data were addressed | NA, 23-24 |
|  |  | (d) If applicable, describe analytical methods taking account of sampling strategy | NA |
|  |  | (e) Describe any sensitivity analyses | 27-28 & Table 5 |
| Results |  |  |  |
| Participants | 13* | (a) Report numbers of individuals at each stage of study—eg numbers potentially eligible, examined for eligibility, confirmed eligible, included in the study, completing follow-up, and analysed | 4 & Figures 1-2 |
|  |  | (b) Give reasons for non-participation at each stage | Figures 1-2 |
|  |  | (c) Consider use of a flow diagram | Figures 1-2 |
| Descriptive data | 14* | (a) Give characteristics of study participants (eg demographic, clinical, social) and information on exposures and potential confounders | Supp. Tables 1 & 2 |
|  |  | (b) Indicate number of participants with missing data for each variable of interest | NA, 24-25 |
| Outcome data | 15* | Report numbers of outcome events or summary measures | 5-8, & Tables 1-5 |
| Main results | 16 | (a) Give unadjusted estimates and, if applicable, confounder-adjusted estimates and their precision (eg, 95% confidence interval). Make clear which confounders were adjusted for and why they were included | 5-8, & Tables 1-5 |
|  |  | (b) Report category boundaries when continuous variables were categorized | Supp. Tables 1 & 2 |
|  |  | (c) If relevant, consider translating estimates of relative risk into absolute risk for a meaningful time period | NA |
| Other analyses | 17 | Report other analyses done—eg analyses of subgroups and interactions, and sensitivity analyses | Supplementary Figure 1 and 2, Tables 2-3 & Table 5 |
| **Discussion** |  |  |  |
| Key results | 18 | Summarise key results with reference to study objectives | 8-9 |
| Limitations | 19 | Discuss limitations of the study, taking into account sources of potential bias or imprecision. Discuss both direction and magnitude of any potential bias | 9-10 |
| Interpretation | 20 | Give a cautious overall interpretation of results considering objectives, limitations, multiplicity of analyses, results from similar studies, and other relevant evidence | 11 |
| Generalisability | 21 | Discuss the generalisability (external validity) of the study results | 10 |
| **Other information** |  |  |  |
| Funding | 22 | Give the source of funding and the role of the funders for the present study and, if applicable, for the original study on which the present article is based | 12 |

Abbreviations: NA, not applicable; Supp, Supplementary Material

